# The psychosocial impact of disability and stigma among persons affected by leprosy and lymphatic filariasis in India

**DOI:** 10.1101/2025.07.04.25330882

**Authors:** Robin van Wijk, Ashok Agarwal, Pradeepta Nayak, Chandra Pati Mishra, Sneha Toppo, Rohit K. Tiwari, Pravin Kumar, Amit Jain, Marente M. Mol, Arup K. Chakrabartty, Jan Hendrik Richardus, Ida J. Korfage, Wim van Brakel

## Abstract

**Background:** Neglected tropical diseases (NTDs), such as leprosy and lymphatic filariasis (LF), can cause disabilities despite available treatment. Both diseases disproportionately affect India’s poorest populations, with high numbers in Jharkhand and Uttar Pradesh. Those affected experience severe psychosocial consequences, including participation restrictions, depression, and poor mental health. This study investigates the extent to which stigma affects social participation and mental wellbeing among persons with leprosy- and LF-related disabilities in two Indian states.

**Methodology:** In a cross-sectional design in Bokaro (Jharkhand) and Jaunpur (Uttar Pradesh), participants were randomly selected among persons with leprosy- or LF-related disabilities, along with comparable non-affected community members. Sociodemographic and disability data were collected, alongside four measures assessing mental wellbeing, depression, participation restriction, and stigma. Analysis used descriptive statistics, univariate, and multivariate regression.

**Results:** A total of 201 persons with leprosy-related disabilities, 240 with LF-related disabilities, and 98 non-affected community members were included. Persons with NTD-related disabilities exhibited significantly higher depression and participation restriction and reported lower mental wellbeing compared to community members (p<0.001). In the leprosy group, lower education and lower stigma were key determinants of mental wellbeing (R²=0.3), while in the LF group, higher age and less severe lymphoedema grade were significant (R²=0.5). Participation restriction was associated with higher age, poorer mental wellbeing, and depression in the leprosy group (R²=0.7), and with depression and stigma in the LF group (R²=0.6).

**Conclusion:** Persons with leprosy and LF-related disabilities face significant psychosocial challenges, including poor mental wellbeing, depression, and restrictions in participation, often exacerbated by stigma. Addressing these challenges requires integration of mental health support and stigma reduction within NTD programmes, with specific attention to the needs of persons with disabilities. Further research into connections between education, disability, and mental health is essential to deepen understanding and inform development of more targeted, effective interventions.

**Key Messages:** *What is already known on this topic:* Leprosy and lymphatic filariasis (LF) are Neglected Tropical Diseases (NTDs) that can cause long-term physical impairments and are linked to stigma, discrimination, and poor mental health. However, the associations between these psychosocial factors have been underexplored and are often not addressed in disease programmes.

*What this study adds:* This study shows that persons affected by leprosy and LF in India experience worse mental wellbeing and greater participation restrictions than community members, with stigma playing a key role. Leprosy-affected participants reported higher overall stigma than those with LF.

*How this study might affect research, practice or policy:* The findings highlight the importance of integrating stigma reduction and mental health support into NTD programmes. Peer-led approaches like Basic Psychological Support for persons affected by NTDs (BPS-N) offer a promising model for improving psychosocial outcomes.

## Background

Neglected tropical diseases (NTDs) are a group of twenty-one infectious diseases caused by various pathogens. (1) The World Health Organization (WHO) estimates that globally over 1 billion people are affected by NTDs, and 1.6 billion people need treatment, prevention, or rehabilitation measures for these diseases. NTDs have several characteristics in common (2,3): They affect the poorest people, especially those without access to clean water and sanitation, and to basic healthcare facilities. Many of the NTDs are chronic or progressive diseases when left untreated, causing irreversible damage, including severe chronic pain and lifelong disabilities ref.

Leprosy and lymphatic filariasis (LF) are both among the group of NTDs (1). Leprosy is caused by the mycobacterium leprae and has a global incidence of more than 200,000 per year ref. Leprosy affects the skin, peripheral nerves and the eyes, leading to insensitive skin patches, loss of sensation and motor function in mainly the hands and feet, and blindness. (4,5) Even though treatment is available in the form of multi-drug therapy (MDT), which is a combination of three antibiotics, diagnosis often comes late, leaving the affected person with irreversible disabilities. (6) LF, also known as elephantiasis, is caused by filarial parasites transmitted by mosquitos. (7) Although some people are asymptomatic carriers of the disease, for about half of the persons with an active infection, LF affects the lymphatic system leading to lymphoedema in the limbs and/or hydrocele (scrotal swelling). (7,8) Persons affected by LF also experience acute attacks of local inflammation, causing acute illness.

India is home to a large percentage of the global leprosy and LF cases. (9) The WHO reported 107,851 new leprosy cases in 2023, accounting for 59% of the global leprosy incidence. (9) The Indian Ministry of Health and Family Welfare reported that in 2022, the leprosy incidence is highest in the states of Bihar, Chhattisgarh and Jharkhand. Uttar Pradesh also accounts for a significant number of new cases. (10) At national level, out of the newly identified persons affected by leprosy, around 2.3% have a grade 2 disability at the time of diagnosis and 5.4% are child cases (less than 15 years of age). (9) For LF, there are estimated to be up to 31 million people infected with microfilariae who are at risk of developing LF, 23 million cases of symptomatic LF, and about 473 million individuals potentially at risk of infection in India. (11) Indigenous LF cases are common in Jharkhand and Uttar Pradesh. (12)

Both leprosy and LF are associated with severe psychosocial consequences. (13–15) The impairments caused by the diseases impede affected persons in their activities, often resulting in participation restriction, for example the limited ability to work. Eventually, this may lead to exclusion from education, work and social gatherings. On top of the physical impairments leading to disability, persons affected by leprosy and LF also often experience stigma and discrimination, frequently connected to the infectious and disfiguring character of the diseases. (13,16) The associated stigma adds to the aforementioned social exclusion. Stigma also has a negative influence on the health-seeking behaviour of patients, which -in turn - leads to prolonged transmission of the disease and worsening of impairments. Related to this, mental wellbeing of persons affected by NTDs tends to be poor. (17,18) Depression, anxiety and suicidal thoughts are frequently present in persons with these diseases, leading to chronic comorbidity and increasing disability. (17,19,20)

Several studies have shown that the psychosocial consequences of leprosy for persons living in India are significant. (21–23) Similar consequences have been shown for persons with LF-related lymphoedema or hydrocele. (24–26) However, associations between the various factors have not been quantified and psychosocial consequences are often poorly addressed in disease control programmes. The present study aimed to examine the extent to which stigma affects social participation and mental wellbeing among persons with leprosy- and LF-related disabilities in Jaunpur in Uttar Pradesh and Bokaro in Jharkhand in India.

## Methodology

### Study Design

This research used a cross-sectional design with quantitative methods. The design allowed analysis of psychosocial outcomes and the influence of disability and stigma of a sample of persons affected by leprosy and LF in the Bokaro (Jharkhand) and Jaunpur (Uttar Pradesh) districts in India. In addition, a sample of community members was included to assess the status of mental wellbeing, depression levels and participation restrictions of community members who are not affected by leprosy or LF. Mental wellbeing scores of the community members were also used to come to context-specific cut-off values for this measurement. The data collection was done between 18 October 2021 and 16 December 2021.

### Study Population

#### Sampling

The study population comprised persons with leprosy or LF-related disabilities, as well as community members in the Bokaro and Jaunpur districts. A purposive sampling strategy was used to select states and districts known for a high prevalence of these diseases. Random sampling was then applied at the block level to select the participants affected by leprosy or LF. In addition, a sample of community members was selected through purposive sampling.

All persons with confirmed diagnoses of leprosy or LF and with related disabilities residing in the districts were eligible. Upon assessment by a trained psychologist, persons with serious mental health problems and illness who could not respond to questions were excluded.

Community Members: based on mapping data, the block with the highest number of affected persons, a community member of (similar gender to the leprosy or LF case) in the vicinity of the leprosy and LF case was selected for the study. Family members, relatives or direct neighbours were excluded.

#### Sample size

To obtain a prevalence estimate with a 95% confidence interval of +/- 10%, a total sample of 100 persons for each disease group in each district was required. This led to an aimed sample size of 200 with leprosy-related disabilities and 200 persons with LF-related disabilities.

The additional study focusing on community members was performed only in Bokaro district. The aimed sample size was 96. This was based on obtaining a precision of +/- 10% of a prevalence estimate of 50% for poor mental wellbeing.

### Data Collection

The data collection included socio-demographic and disability data. For leprosy-related disability, the WHO Disability Grading system (0–2) was applied to assess the severity of impairment in the eyes, hands, and feet, using the Eye, Hand, Foot (EHF) score. (27) Each of these body parts was graded from 0 to 2 based on the severity of impairment. The highest grade for any of these areas represented the ‘maximum disability grade’ for the person affected. For LF-related disability, the WHO grading system for lymphoedema was utilised. (28) This system assigns a grade based on the severity of oedema: Grade 1 indicates mostly pitting oedema that is reversible on elevation of the leg; Grade 2 indicates mostly non-pitting oedema that is not reversible on elevation of the leg; and Grade 3, or elephantiasis, is characterised by gross swelling of the limb, non-pitting oedema, thickened skin, and irreversibility on elevation of the leg.

Four standardised measures were used to assess participants’ levels of mental wellbeing, depression, participation restriction and stigma, respectively:

1. The Warwick-Edinburgh Mental Wellbeing Scale (WEMWBS) has 14 items with response options ranging from 1 (none of the time) to 5 (all the time), resulting in a score range of 14-70 (29) Cut-off scores for low, normal or high mental wellbeing were determined through the community sample included in this study.
2. The Patient Health Questionnaire 9 items (PHQ-9) has 9 items with response options ranging from 0 (not at all) to 3 (nearly every day), resulting in a score range of 0-27. Threshold scores indicate no (0-4) mild (5-9), moderate (10-14), moderately severe (15-19) and severe depression (20-27), respectively. (30)
3. Participation Scale Short Simplified (PSSS) has 13 items with response options ranging from 0 (easy) to 4 (very difficult), resulting in a score range of 0-52. Threshold scores indicate no (0-7); mild (8-13); moderate (14-20); and severe restrictions (21-52). (31)
4. The SARI Stigma Scale (SSS) assesses four stigma domains (experienced, internalized and perceived stigma, and disclosure concerns) and has 21 items. The response options are no (0), rarely (1), sometimes (2) and always/often (3), resulting in a score range of 0-63. Higher scores indicate higher stigma levels. (32)

The SSS and PSSS were validated in Hindi during the formative study, while Hindi versions of the PHQ-9 and WEMWBS had already been validated in previous studies. (33,34,35)

### Data Analysis

Data analysis was performed using SPSS v27, with both descriptive and inferential statistical methods applied. Missing values for questionnaire items were imputed by the study participant’s average score on the scale. Descriptive statistics such as means, medians, frequencies, and percentages were used to summarise socio-demographic data. For normally distributed data, two-sample t-tests and one-way ANOVA with post-hoc Tukey’s tests were employed to compare mental wellbeing, depression, participation and stigma scores across subgroups. Non-normally distributed data were analysed using the Kruskal-Wallis test, with pairwise Mann-Whitney U tests for post-hoc comparisons. Based on a sample of 98 community controls, cut-offs for low mental wellbeing (bottom 15%), average mental wellbeing (16-85%) and high mental wellbeing (top 15%) were chosen, as was done in the original study from the UK that developed the WEMWBS measure. (29)

A multivariate linear regression analysis was conducted to explore associations between demographic variables and key outcomes. Bootstrapping was used to account for non-normal data distributions, and independent variables with a p-value < 0.2 in univariate analysis were included in the model. Variables were removed stepwise until only significant predictors (p < 0.05) remained.

### Ethical Considerations

Ethical approval for the study was granted by Banaras Hindu University’s Ethics Committee (No. Dean/2020/EC/882), and the study also obtained clearance from the India Health Ministry’s Screening Committee. All participants provided informed consent, and confidentiality and anonymity were maintained throughout the study.

## Results

In total, 539 participants were included in the study. These comprised 201 persons affected by leprosy, of which 102 were male participants and 99 were female participants. Of the total 240 persons affected by LF included in the study, 112 were male particpants and 128 were female participants. The mean age was 46 and 52 years for the leprosy and LF group, respectively. All leprosy and LF-affected participants had at least one form of disability. 58% of the persons affected by leprosy had grade 1 disabilities, while 42% had grade 2 disabilities. For persons affected by LF, 48% had grade 1 disabilities, 39% had grade 2 disabilities and 13% had grade 3 disabilities. A total of 98 community members were included, of which 63 were male participants and 35 were female participants. Their mean age was 46 years. Table 1 provides the full overview of study participant characteristics.

**Table 1:** Characteristics of the study sample.

|  |  | Persons affected by leprosy |  | Persons affected by lymphatic filariasis |  | Community members |  |
| --- | --- | --- | --- | --- | --- | --- | --- |
|  |  | n | % | n | % | n | % |
| **Total** |  | 201 |  | 240 |  | 98 |  |
| **Sex** | Male | 102 | 50.7 | 112 | 46.7 | 63 | 64.3 |
|  | Female | 99 | 49.3 | 128 | 53.3 | 35 | 35.7 |
| **Age (years)** | Mean (SD) | 46.4 (18.8) |  | 52.7 (15.5) |  | 46.4 (12.1) |  |
| **District of residence** | Jaunpur | 103 | 51.2 | 115 | 47.9 | 0 | 0 |
|  | Bokaro | 98 | 48.8 | 125 | 52.1 | 98 | 100 |
| **Marital status** | Married | 140 | 69.7 | 191 | 79.6 | 95 | 96.9 |
|  | Single | 32 | 15.9 | 14 | 5.8 | 1 | 1.0 |
|  | Widowed | 28 | 13.9 | 33 | 13.8 | 2 | 2.1 |
|  | Divorced | 1 | 0.5 | 2 | 0.8 | 0 | 0 |
| **Education level** | None | 109 | 54.2 | 130 | 54.2 | 24 | 24.5 |
|  | Primary | 33 | 16.4 | 38 | 15.8 | 14 | 14.3 |
|  | Middle | 24 | 11.9 | 27 | 11.3 | 18 | 18.3 |
|  | Secondary | 18 | 9 | 14 | 5.8 | 16 | 16.3 |
|  | Higher secondary | 9 | 4.5 | 22 | 9.2 | 17 | 17.3 |
|  | Tertiary | 8 | 4.0 | 9 | 3.8 | 9 | 9.2 |
| **Disability/ lymphoedema grade** | 0 | 0 | 0 | 0 | 0 | n/a |  |
|  | 1 | 116 | 57.7 | 114 | 47.5 | n/a |  |
|  | 2 | 85 | 42.3 | 94 | 39.2 | n/a |  |
|  | 3 | N/a |  | 32 | 13.3 | n/a |  |

### Mental wellbeing, depression and participation restriction

A mean score of 32.5 and 30.9 for mental wellbeing on the WEMWBS was found for persons affected by leprosy and LF, respectively. For community members this was 36.3, which is significantly higher (p<0.001). When looking at the PHQ-9 scores for depression, persons affected by leprosy had a mean score of 9.0 and persons affected by LF a mean score of 8.7. For community members the mean depression score is 3.6, which is significantly lower (p<0.001). Median scores found on the PSSS for both persons affected by leprosy and LF are 12. For community members this was 6.5. A complete overview of questionnaire scores per study group can be found in Table 2.

**Table 2:**
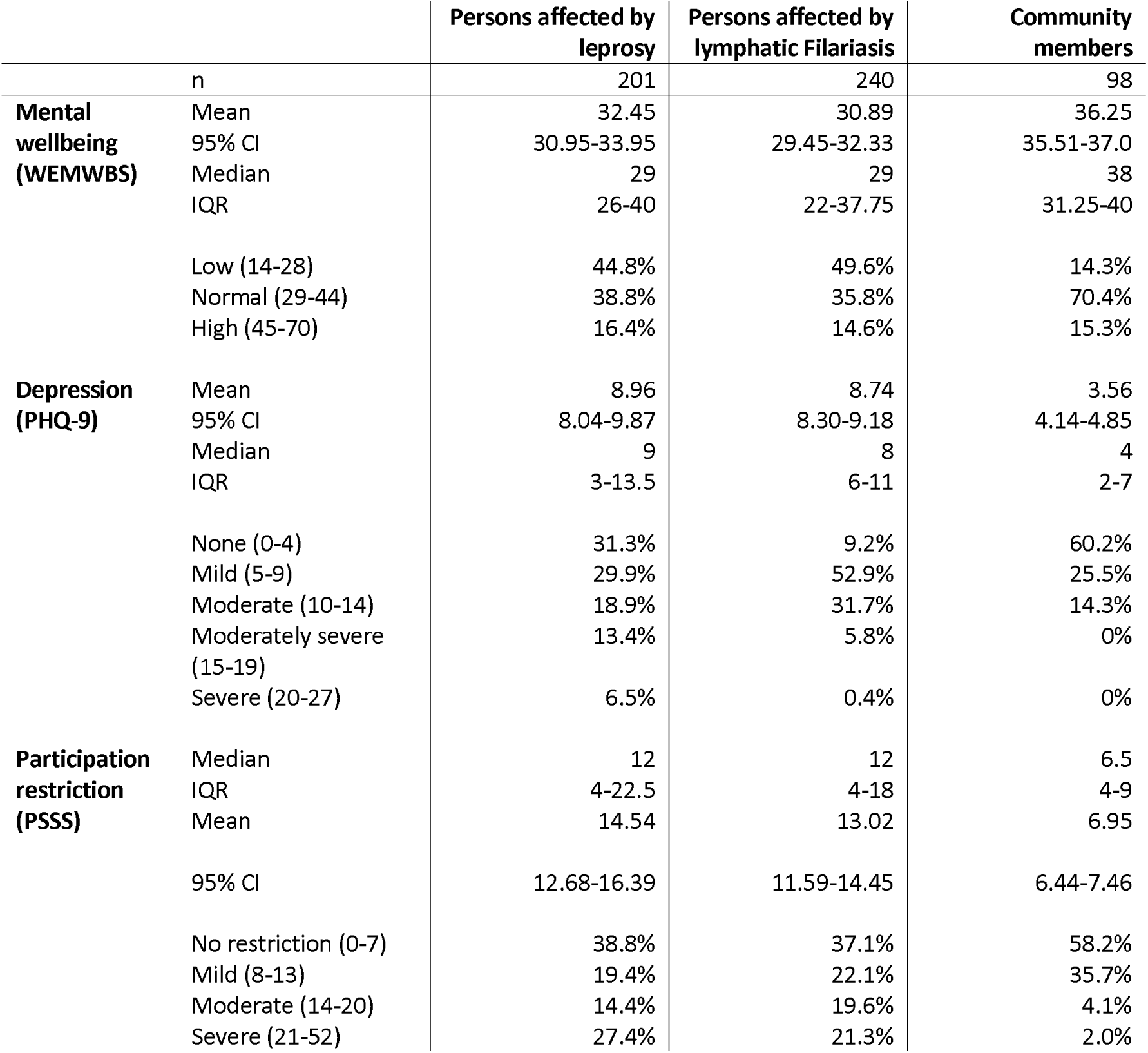
Overview of questionnaire outcomes for mental wellbeing, depression and participation restriction.

### Stigma

Both groups of persons affected by leprosy and LF reported stigma on the SSS. Median scores were 13 (IQR: 5-32) and 6 (IQR: 2-12), respectively. A complete overview of stigma scores can be found in Table 3. A significant difference in total stigma scores between disability grades was not found for persons affected by leprosy (p=0.069). For persons affected by LF, a significant difference in stigma scores was found between lymphoedema grade 3 and lower lymphoedema grades (p=0.002). Figure 1 shows the distribution of stigma scores per disability and lymphoedema grade.

**Figure 1:**
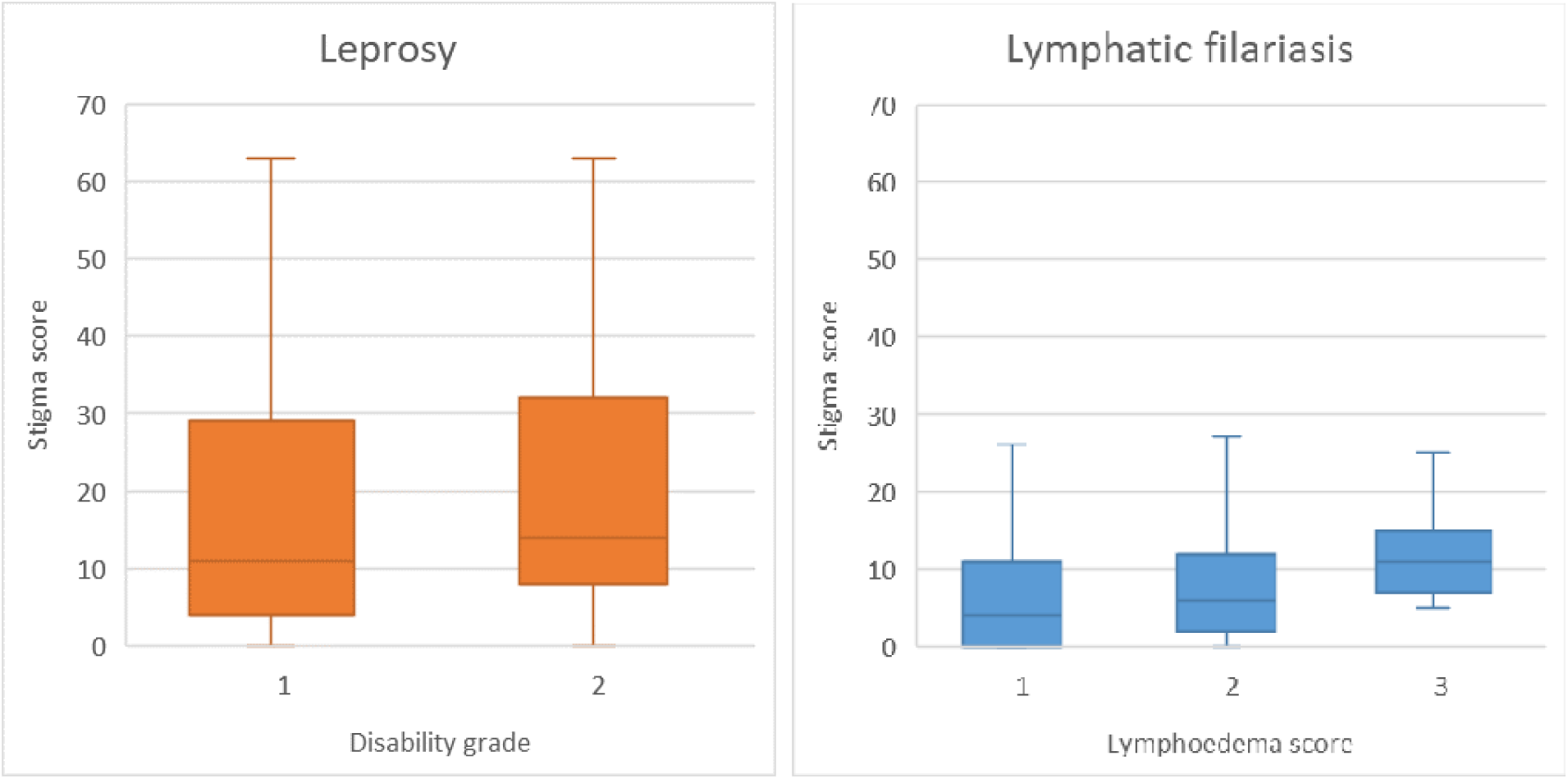
Stigma scores per disability/lymphoedema grade Notes: IQR = Interquartile range; 95% CI = 95% confidence interval

**Figure 2:**
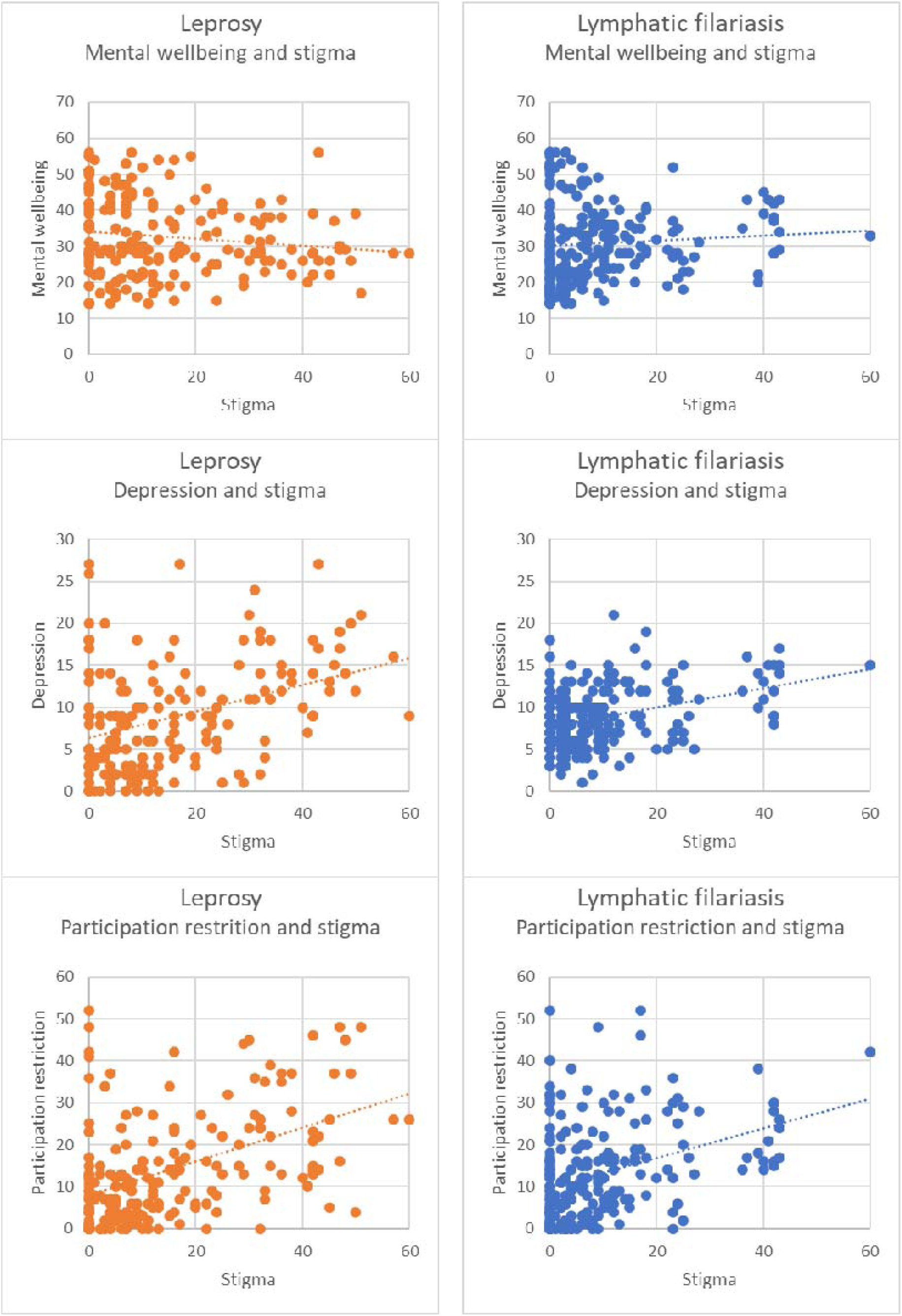
Associations between stigma and psychosocial outcomes among persons affected by leprosy and LF. Each dot represents one participant. The x-axis shows individual stigma scores (SSS), while the y-axis reflects scores for mental wellbeing (WEMWBS), depression (PHQ-9), or participation restriction (PSSS). Trend lines indicate the direction and strength of the associations across the six graphs.

**Table 3:** Questionnaire outcomes for stigma measurement.

|  |  | Persons affected by leprosy | Persons affected by Lymphatic Filariasis |
| --- | --- | --- | --- |
|  | n | 201 | 240 |
| **SARI Stigma Scale** | Median | 13 | 6 |
|  | IQR | 5-32 | 2-12 |
|  | Mean | 18.6 | 9.75 |
|  | 95% CI | 16.2-21.2 | 8.21-11.3 |
| **Experienced stigma** |  |  |  |
|  | Mean score per question | 0.91 | 0.31 |
|  | 95% CI | 0.77-1.1 | 0.23-0.39 |
| **Disclosure concern** |  |  |  |
|  | Mean score per question | 0.88 | 0.71 |
|  | 95% CI | 0.76-1.0 | 0.61-0.81 |
| **Internalised stigma** |  |  |  |
|  | Mean score per question | 0.85 | 0.61 |
|  | 95% CI | 0.73-0.97 | 0.53-0.70 |
| **Anticipated stigma** |  |  |  |
|  | Mean score per question | 0.93 | 0.44 |
|  | 95% CI | 0.81-1.1 | 0.35-0.52 |

### Factors influencing psychosocial wellbeing

The outcomes of univariate analysis for associations between mental wellbeing, depression and participation restriction, and sociodemographic factors are shown in Table 4. Associations between the psychosocial outcomes and stigma are visualised in Figure 1.

**Table 4:**
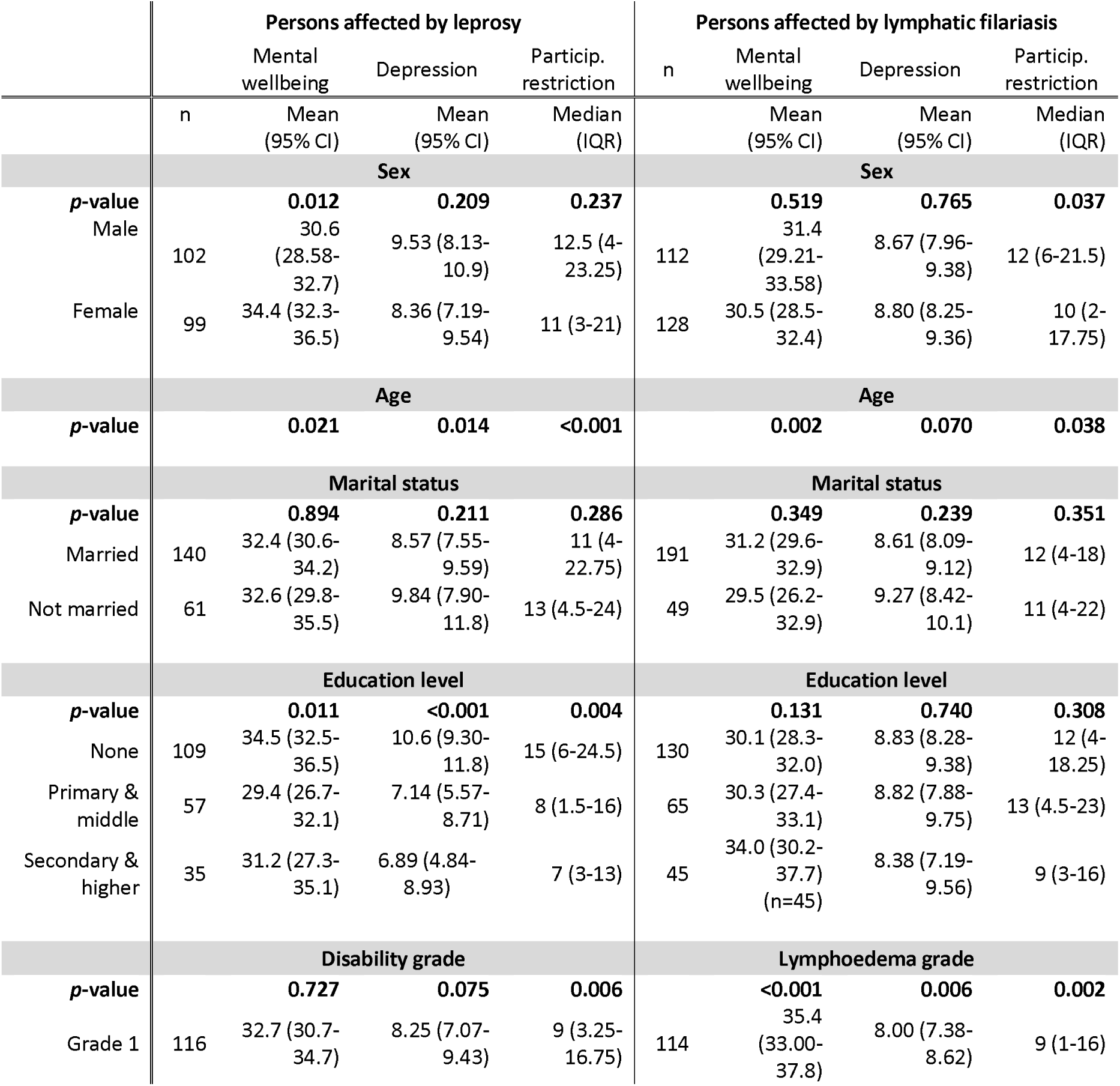
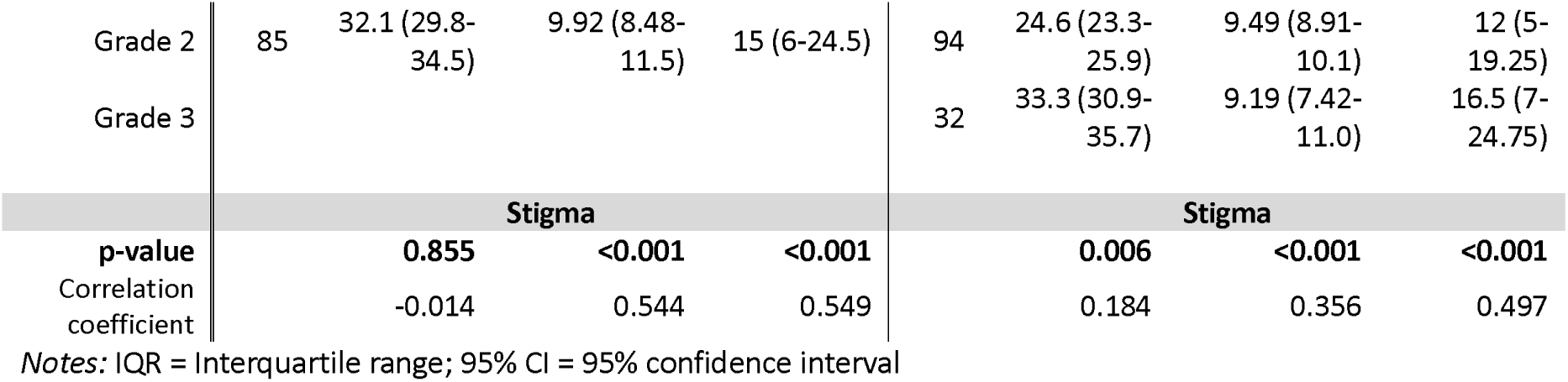
Sociodemographic factors associated with mental wellbeing, depression and participation restriction.

### Persons affected by Leprosy

Significant associations for persons affected by leprosy were found for mental wellbeing and sex (p=0.012), age (p=0.021) and education level (p=0.011). For depression, significant associations were found with age (p=0.014) and education level (p<0.001). And for participation restriction, significant associations were found with age (p<0.001), education level (p=0.004) and disability grade (p=0.006). We found associations between stigma and depression (p<0.001) and stigma and participation restriction (p<0.001).

### Persons affected by lymphatic filariasis

Significant associations for persons affected by LF were found for mental wellbeing and age (p=0.0201) and lymphoedema grade (p<0.001). For depression significant associations were found with lymphoedema grade (p=0.006). And for participation restriction significant associations were found with sex (p=0.037), age (p=0.038), and lymphoedema grade (p=0.002). Similar for persons affected by leprosy we also found associations between stigma and depression (p<0.001) and stigma and participation restriction (p<0.001) for persons affected by LF.

### Stigma and other determinants related to psychosocial wellbeing

Negative associations between stigma and depression, and stigma and participation restriction in both disease groups were observed (Table 5). Especially for participation restriction, a strong association between stigma and depression was found, explaining up to 59% of the variance in persons affected by leprosy. For persons affected by leprosy, higher education is associated with poorer mental wellbeing and for persons affected by LF, multivariate analysis shows that older age and a higher lymphoedema grade contribute to poorer mental wellbeing. When looking at the combined sample, consisting of both persons affected by leprosy and LF, we note that both a visible disability and education are determinants for poorer mental wellbeing and depression.

**Table 5:**
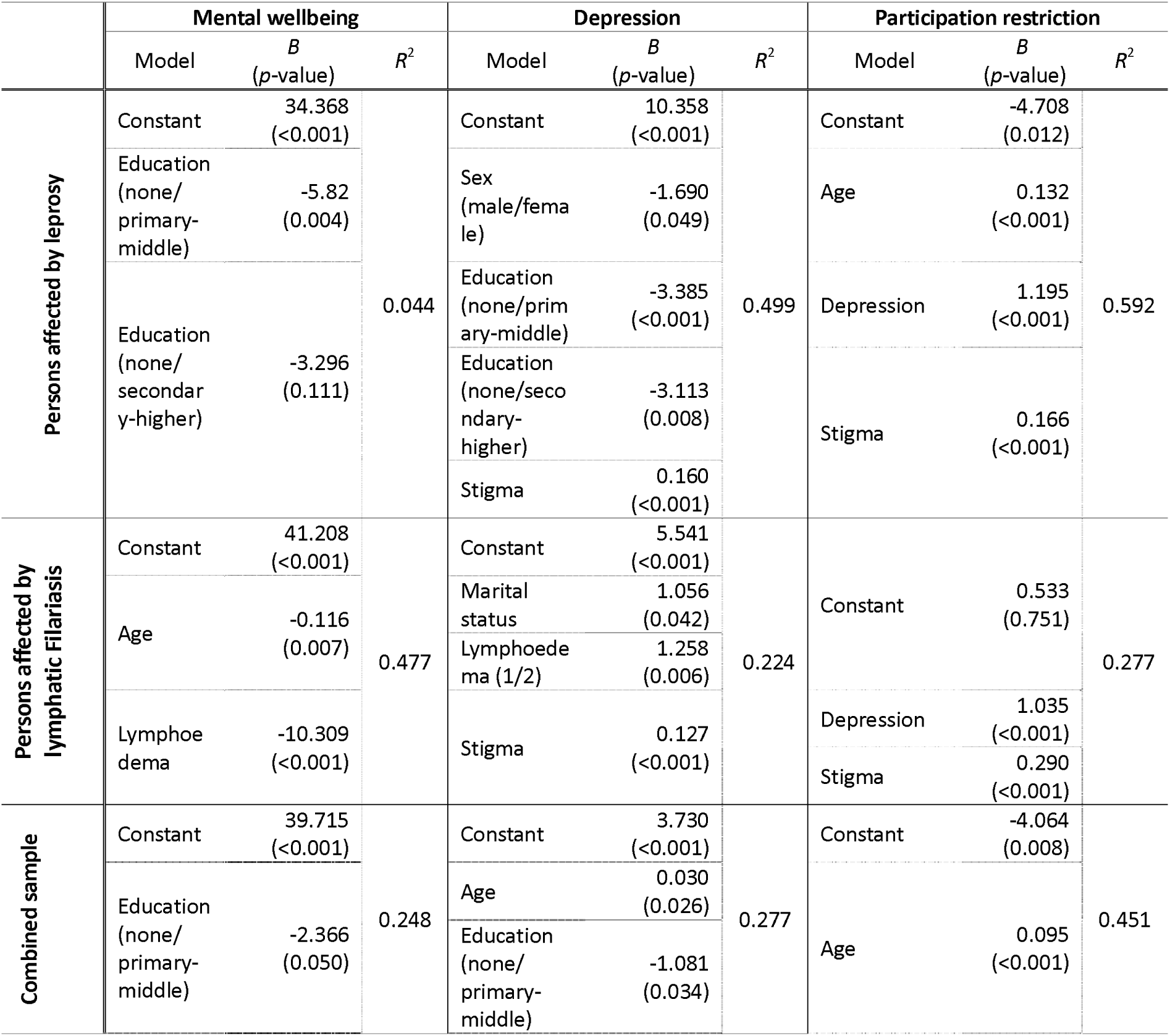
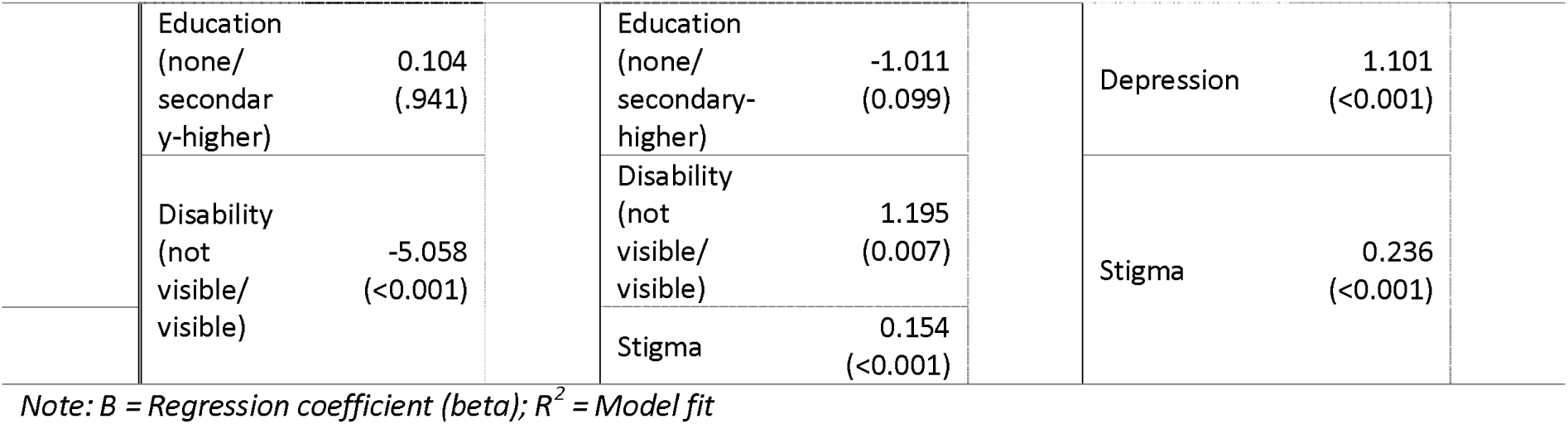
Multivariate models for mental wellbeing, depression and participation restriction in persons affected by leprosy and LF, and combined.

|  | Mental wellbeing |  |  | Depression |  |  | Participation restriction |  |  |
| --- | --- | --- | --- | --- | --- | --- | --- | --- | --- |
|  | Model | B<br>(p-value) | R<sup>2</sup> | Model | B<br>(p-value) | R<sup>2</sup> | Model | B<br>(p-value) | R<sup>2</sup> |
| Persons affected by leprosy | Constant | 34.368<br>($<0.001$) | 0.044 | Constant | 10.358<br>($<0.001$) | 0.499 | Constant | -4.708<br>(0.012) | 0.592 |
| | Education<br>(none/<br>primary-<br>middle) | -5.82<br>(0.004) | | Sex<br>(male/female) | -1.690<br>(0.049) | | Age | 0.132<br>($<0.001$) | |
| | Education<br>(none/<br>secondary-<br>higher) | -3.296<br>(0.111) | | Education<br>(none/primary-<br>middle) | -3.385<br>($<0.001$) | | Depression | 1.195<br>($<0.001$) | |
| | | | | Education<br>(none/secondary-<br>higher) | -3.113<br>(0.008) | | Stigma | 0.166<br>($<0.001$) | |
| | | | | Stigma | 0.160<br>($<0.001$) | | | | |
| Persons affected by lymphatic Filariasis | Constant | 41.208<br>($<0.001$) | 0.477 | Constant | 5.541<br>($<0.001$) | 0.224 | Constant | 0.533<br>(0.751) | 0.277 |
| | Age | -0.116<br>(0.007) | | Marital<br>status | 1.056<br>(0.042) | | Depression | 1.035<br>($<0.001$) | |
| | Lymphoedema | -10.309<br>($<0.001$) | | Lymphoedema (1/2) | 1.258<br>(0.006) | | Stigma | 0.290<br>($<0.001$) | |
| | | | | Stigma | 0.127<br>($<0.001$) | | | | |
| Combined sample | Constant | 39.715<br>($<0.001$) | 0.248 | Constant | 3.730<br>($<0.001$) | 0.277 | Constant | -4.064<br>(0.008) | 0.451 |
| | Education<br>(none/<br>primary-<br>middle) | -2.366<br>(0.050) | | Age | 0.030<br>(0.026) | | Age | 0.095<br>($<0.001$) | |
|  |  |  |  | Education<br>(none/<br>primary-<br>middle) | -1.081<br>(0.034) |  |  |  |  |

|  |  |  |  |  |  |  |  |  |
| --- | --- | --- | --- | --- | --- | --- | --- | --- |
|  | Education<br>(none/<br>secondary-<br>higher) | 0.104<br>(.941) |  | Education<br>(none/<br>secondary-<br>higher) | -1.011<br>(0.099) |  | Depression | 1.101<br>(<0.001) |
|  | Disability<br>(not<br>visible/<br>visible) | -5.058<br>(<0.001) |  | Disability<br>(not<br>visible/<br>visible) | 1.195<br>(0.007) |  | Stigma | 0.236<br>(<0.001) |
|  |  |  |  | Stigma | 0.154<br>(<0.001) |  |  |  |

Note: B = Regression coefficient (beta); $R^2$ = Model fit

## Discussion

The present study aimed to examine the extent to which stigma affects social participation and mental wellbeing among persons with leprosy- and LF-related disabilities in Jaunpur and Bokaro districts in India. The study found notable differences in mental wellbeing, depression, and stigma between persons affected by leprosy and LF, and community members. Those affected by leprosy and LF reported worse mental wellbeing scores and more depression compared to community members. Both groups were confronted with stigma, but participants affected by leprosy experienced higher levels of stigma than those affected by LF. Among the LF group, higher scores were found for disclosure concerns and internalised stigma. Additionally, significant associations were identified between stigma, depression, and participation restriction, highlighting the negative impact of these factors on their overall wellbeing and social inclusion.

The presence of physical disabilities resulting from leprosy or LF influences the psychosocial wellbeing of affected persons, underscoring the complex interplay between disability, stigma, mental health, and social participation. All participants in the study sample of persons affected by leprosy and LF had disease-related physical disabilities. A scoping review by Moreira de Araujo et al. examined the impact of physical disability caused by leprosy on the quality of life of affected persons, reporting negative effects in areas such as activities of daily living, social and economic engagement, and mental wellbeing. (35) Similarly, a systematic review by Ali et al., which included studies on both leprosy and LF, found “a significant association between being affected by the selected NTD and psychosocial and mental health outcomes” (p.13) across varying degrees of disability. (36) Aggregated analysis across both disease groups revealed a significant association between visible disabilities and poorer mental wellbeing and depression. This suggests that visible disabilities among persons affected by NTDs exacerbate psychosocial challenges. These findings align with a recent study by Belgaumkar et al., which reported that the quality of life is poorer for persons affected by leprosy with visible disabilities than for those without. (37) Anand et al., who investigated the quality of life of individuals with severe leprosy and LF-related disabilities in India, noted that “data analysis shows that interventions addressing urgent physical needs have the potential to positively impact the quality of life of up to 90% of the people with severe NTD disability” (p.13), highlighting the critical role of addressing physical impairments to improve wellbeing. (38)

Our results showed that persons affected by leprosy and LF experienced significantly poorer mental wellbeing, higher depression levels, and greater participation restrictions compared to community members, with stigma playing a central role in exacerbating these outcomes. Somar, Waltz and van Brakel conducted a systematic review to consolidate evidence regarding the mental health impact of leprosy. (39) They described that depressive and anxiety disorders are common among persons affected by leprosy. They also confirmed that stigma and discrimination are frequently associated with these outcomes. They made note of the link between visible impairments and mental health outcomes. Koschorke et al.(41) were able to confirm through an extensive review that the prevalence of mental health conditions among persons affected by NTDs was higher than in the general population. (40) In their review, Alderton et al. described the psychosocial impacts of NTDs as perceived by the persons affected. (41) They made specific note of the participation restrictions faced by persons affected by LF related to the severity of their disability. These were not only related to activity limitations, but also because of social exclusion in their communities and difficulties in marriage or finding a marriage partner. (24,25,41,42)

In the current study, mental wellbeing for persons affected by leprosy seems better for those without any education than for those with primary, secondary or tertiary level education, which is not in line with common findings in literature. This is interesting, when compared to outcomes of the Indian National Mental Health survey conducted among 34,082 participants, which clearly shows an increased risk of poor mental wellbeing for those with lower or no education levels. (43) In line with these findings in the general population, Govindsamy et al. studied depression and anxiety among persons affected by leprosy and also found these were more common among those with lower education levels. (44) A follow-up study with a larger sample size and qualitative components would therefore be recommended to confirm the effect of education level on mental wellbeing and to find explanations for this finding.

While the presence of disability and stigma may lead to poorer mental wellbeing, depression and participation restriction, interventions that provide psychosocial support can help mitigate these effects. Basic Psychological Support for persons affected by NTDs (BPS-N) as used by Agarwal et al. directly addresses the challenges of physical disabilities, stigma, mental health issues, and participation restrictions experienced by persons affected by leprosy and LF. (45) Peer support, an essential element of the BPS-N approach, has shown to be particularly effective in promoting connectedness, fostering hope, and offering a unique level of understanding that is often absent in professional services. (46,47) In addition, Lusli et al. reported that counselling sessions facilitated by peer counsellors demonstrate the value of shared experiences in building trust and encouraging reflection. (48) Many participants valued the personal insights shared by their counsellors, which strengthened the impact of the intervention. The BPS-N intervention in combination with peer-counselling can potentially provide a vital framework for improving the wellbeing of persons affected by NTDs through community-based and person-centred support. (46,47)

A key strength of the study is that the cross-sectional design, combined with quantitative methods, enabled a broad snapshot of psychosocial outcomes and their associations with disability and stigma among persons affected by leprosy and LF. While causal relationships cannot be determined, the design allowed for valuable comparisons across groups and identification of significant patterns in mental wellbeing, depression, participation, and stigma. Including a sample of community members provided a valuable baseline to contextualise disease-specific impacts and strengthened the study’s validity. Four standardised questionnaires, WEMWBS, PHQ-9, PSSS, and SSS, were used to assess mental wellbeing, depression, participation restriction, and stigma. These tools, validated in Hindi, ensured cultural and linguistic appropriateness. Additionally, cut-off values for the WEMWBS were adapted using data from the community sample, enabling meaningful, context-specific comparisons. This study has several limitations. Some subgroup analyses, such as by gender or disability severity, may have lacked sufficient statistical power due to sample size constraints, potentially masking important differences. The cut-off scores for mental wellbeing were derived from a local community sample and may not be applicable to other populations, limiting comparability with other studies.

While Hindi versions of the scales were properly translated and validated, not all participants spoke Hindi. As a result, data collectors had to make on-the-spot translations, which may have affected the consistency and reliability of responses.

## Conclusion

Persons with leprosy and LF-related disabilities face significant psychosocial challenges, including poor mental wellbeing, depression, and participation restrictions, challenges that are not solely the result of physical impairments, but are closely linked to the experience of stigma. Addressing these issues requires integrating mental health support and stigma reduction into NTD programmes, with particular attention to persons with disabilities. The BPS-N intervention, especially when combined with peer counselling, can potentially provide a vital framework for improving the wellbeing of persons affected by NTDs through community-based and person-centred support. Further research into the connections between education, disability, and mental health is essential to deepen understanding of these complex intersections and to guide the development of more effective, inclusive interventions.

## List of Abbreviations

BPS-N: Basic Psychological Support for persons affected by NTDs
CI: Confidence Interval
COR-NTD: Coalition for Operational Research on Neglected Tropical Diseases
EHF score: Eye, Hand, Foot score
IQR: Interquartile Range
LF: Lymphatic Filariasis
MDT: Multi-Drug Therapy
NTDs: Neglected Tropical Diseases
PHQ-9: Patient Health Questionnaire, 9 items
PSSS: Participation Scale Short Simplified
SD: Standard Deviation
SSS: SARI Stigma Scale
WEMWBS: Warwick-Edinburgh Mental Wellbeing Scale
WHO: World Health Organization

## Data Availability

All data produced are available online at

https://data.mendeley.com/datasets/bp82sxcsz8/1

## Acknowledgements

The authors sincerely thank the Indian government authorities of Bokaro (Jharkhand) and Jaunpur (Uttar Pradesh), as well as the state governments of Jharkhand and Uttar Pradesh and the Central Government, for their invaluable support. Our heartfelt gratitude also goes to the study participants for their time and insights, the peer supporters for their dedication, and the Indian Institute of Development Management for their invaluable assistance with the data collection. The authors acknowledge the use of ChatGPT, an AI language model developed by OpenAI, for language checking and refinement of this article.

## Consent for publication

Not applicable.

## Availability of data and materials

The dataset supporting the findings of this study is openly available and can be accessed at https://data.mendeley.com/datasets/bp82sxcsz8/1. All data have been anonymised to protect participant confidentiality. Access is provided under the terms and conditions specified in the data repository.

## Authors’ contributions

Conceptualization: AA, PN, CPM, WvB; Data curation: PN, ST, RKT; Formal analysis: RvW, AJ; Investigation: RvW, AA, PN, WvB; Methodology: PN, CPM, WvB; Project administration: PN, ST, RKT, PK; Supervision: AA, CPM, JR, IJK, WvB; Validation: RvW, PN, AJ, MM; Visualization: RvW, AJ; Writing – original draft: RvW; Writing – review & editing: RvW, AA, PN, CPM, ST, RKT, PK, AJ, MM, AKC, JR IJK, WvB

